# Case fatality risk by age from COVID-19 in a high testing setting in Latin America: Chile, March-May, 2020

**DOI:** 10.1101/2020.05.25.20112904

**Authors:** Eduardo A. Undurraga, Gerardo Chowell, Kenji Mizumoto

## Abstract

**Background:** Early severity estimates of COVID-19 are critically needed to better assess the potential impact of the ongoing pandemic in different socio-demographic groups. Using real-time epidemiological data from Chile, the nation in Latin America with the highest testing rate for COVID-19, we derive delay-adjusted severity estimates by age group as of May 18^th^, 2020.

**Methods:** We employed statistical methods and daily series of age-stratified COVID-19 cases and deaths reported in Chile to estimate the delay-adjusted case fatality rate across six age groups.

**Results:** Our most recent estimates of the time-delay adjusted case fatality rate are 0.08% (95% Credible Interval CrI:0.04-0.13%) among persons aged 0-39, 0.61% (95%CrI:0.41-0.87%) for those aged 40-49, 1.06% (95%CrI:0.76-1.40%) for those aged 50-59, 3.79% (95%CrI:3.04-4.66%) for those aged 60-69, 12.22% (95%CrI:10.40-14.38%) for those aged 70-79, and 26.27% (95%CrI:22.95-2980%) for persons aged 80 and over. The overall time-delay adjusted case fatality rate is1.78% (95%CrI: 1.63-1.95%) across all age groups.

**Conclusions:** Severity estimates from COVID-19 in Chile indicate a disproportionate impact among seniors, especially among those aged ≥ 70 years. COVID-19 is imposing a high death toll in Latin America. Case fatality rates in Chile suggest the health system is not yet overwhelmed, but the epidemic is expanding fast.

## Background

The coronavirus disease 2019 (CoVID-19) pandemic has strained or overwhelmed health systems across the world [1, 2], with almost five million lab-confirmed cases and 320 thousand deaths as of May 18, 2020 [3, 4]. The first case in Latin America of Severe Acute Respiratory Syndrome Coronavirus 2 (SARS-CoV-2), the cause of CoVID-19, was reported in February 25 in São Paulo, Brazil, a travel hub in the region [5]. A few weeks later, countries in the region had imposed major epidemic control measures, such as closed borders, restricted travel, closure of schools and universities, and enforced lockdowns [6]. Despite these measures, the ongoing coronavirus pandemic has already imposed a high toll to most countries in Latin America, killing thousands in Brazil (21,408), Ecuador (3,056), and Peru (3,244) thus far [3]. In addition to already strained healthcare systems, other factors have affected the dynamic of the pandemic in the region, including migration, sociopolitical crises, struggling economies, other infectious disease outbreaks, and challenges tied to the implementation of social distancing, hygiene, and lockdown strategies, due to inadequate water and sanitation infrastructure, precarious living conditions [6-11].

Chile has seen a rapid increase in CoVID-19 cases in recent weeks, with 46,048 cases and 544 deaths [12] as of May 18, 2020. The first CoVID-19 case in Chile was reported in March 3, 2020 [12]. The Ministry of Health put in place a quick response effort, announcing restrictions on large gatherings in March 13^th^, and subsequently, closing all daycares, schools, and universities (March 16^th^), imposing border controls and telework recommendations (March 18^th^), closing non-essential businesses (March 19^th^), national night curfews (March 22^nd^), and implementing a strategy of intermittent lockdowns in selected municipalities since March 22^nd^ [13-15]. Estimates for the early stage of the epidemic showed sustained transmission with an estimated reproduction number R~1.6 (95% CI: 1.5, 1.7), and 20-day ahead forecast suggested containment measures significantly slowed down the spread of the virus [16]. The crude case fatality rate (CFR), the number of cumulative deaths over the number of cumulative cases, in Chile (1.2%) has remained well below the global average (6.5%) [4, 12]. But in light of limited health system capacity, infectious disease experts have warned about authorities’ excessive confidence over early successes [17]. In fact, models have estimated the number of ill patients could overwhelm treatment capacity late May to mid-June [18], which has not yet occurred.

SARS-CoV-2 infection can result in a wide spectrum of clinical outcomes, including asymptomatic infection, mild symptoms, hospitalization, or death [19-21]. The case fatality rate (CFR) is a commonly used estimate of the severity of an epidemic [22], as it provides a reliable benchmark for public health officials to make decisions about intensity and duration of interventions to mitigate or suppress an epidemic [23]. But obtaining CFR estimates during the course of an epidemic is challenging, as CFR is typically affected by right censoring and ascertainment bias [24-28]. Right-censoring occurs because of a time delay between onset of symptoms and death and may lead to underestimation of CFR; under ascertainment of cases occurs because mild or asymptomatic COVID-19 cases often go undetected by disease surveillance systems, which are not designed to detect all infections [25, 29-31].

Here we provide real-time estimates of adjusted age-specific CFR during the CoVID-19 epidemic in Chile, March through May 2020, to gauge the severity of the SARS-CoV-2 epidemic. In particular, Chile has tested at a higher rate (23.13 total tests per 1000 people by May 22) than any other country in Latin America [32, 33]. The health system has not yet reached its maximum capacity, with 85% and 96% of critical beds in use nationally and in the capital Santiago [34], respectively, so our estimates are still not affected by excess deaths due to healthcare demand exceeding capacity. To our knowledge, these are the first estimates of CFR in Latin America, which could inform critical decisions by public health officials.

## Methods

### Data Sources

We obtained daily cumulative numbers of reported laboratory confirmed COVID-19 cases and deaths stratified by age group from March 3, 2020, through May 18, 2020, from the Ministry of Health [12]. Data on deaths by age group were missing for a few days (March 29 and 31, April 1-3, 6, 8).

### Statistical analysis

For the estimation of CFR in real time, we employed the delay from hospitalization to death, *h_s_*, which is assumed to be given by *h_s_ = H*(*s*) *− H*(*s-1*) for *s>0* where *H*(*s*) is a cumulative density function of the delay from hospitalization to death and follows a gamma distribution with mean 10.1 days and standard deviation (SD) 5.4 days [35]. Let π*_a_,_ti_* be the time-delay adjusted case fatality ratio on reported day ti in area a, the likelihood function of the estimate π*_a,ti_* is

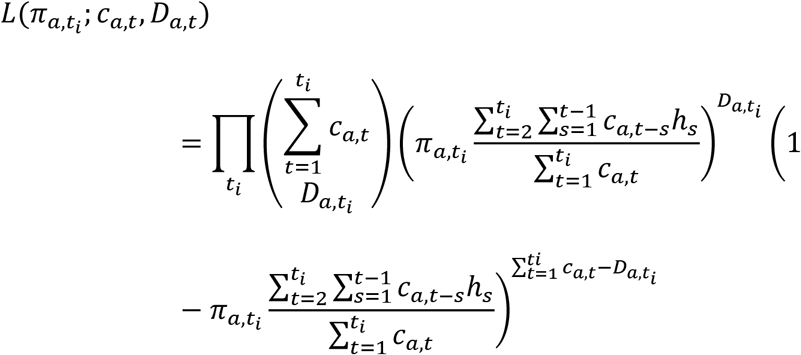

where *c_a_,_t_* represents the number of new cases with reported day *t* in area *a*, and *D_a_,_ti_* is the cumulative number of deaths until reported day t*_i_* in area *a* [36, 37]. Among the cumulative cases with reported day *t* in area *a, D_a_,_ti_* have died and the remainder have survived the infection. The contribution of those who have died with biased death risk is shown in the middle parenthetical term and the contribution of survivors is presented in the right parenthetical term. We assume that *D_a_,_ti_* is the result of the binomial sampling process with probability π*_a,ti_*.

We estimated model parameters using a Monte Carlo Markov Chain (MCMC) method in a Bayesian framework. Convergence of MCMC chains were evaluated using the potential scale reduction statistic [38, 39]. Estimates and 95% credibility intervals (CrI) for these estimates are based on the posterior probability distribution of each parameter and based on the samples drawn from the posterior distributions. All statistical analyses were conducted in R version 3.6.1 (R Foundation for Statistical Computing, Vienna, Austria) using the ‘rstan’ package.

## Results

As of May 18, a total of 46,048 COVID-19 cases and 544 deaths have been reported in Chile. Reported cases were mostly observed among persons aged 0-39 years (53.9%), followed by 40-49 year olds (17.3%), and 50-59 year olds (14.4%) (Table 1). Most reported deaths correspond to seniors, especially those aged 80 years and older (38.1%), followed 70-79 year olds (27.9%), and 60-69 year olds (16.9%) (Table 1).

**Table 1.** Distribution of the CoVID-19 cases in Chile by sex and age groups, as of May 18, 2020

|  |  | Confirmed cases (%) | Deaths (%) | Crude fatality rate (%) |
| --- | --- | --- | --- | --- |
| Total |  | 46,048 (100.00) | 544 (100.00) | 1.18% |
| Sex | Male | 24,523 (53.3) | NA | NA |
|  | Female | 21,525 (46.7) | NA | NA |
| Age-group | 0-39 | 24,830 (53.9) | 11 (2.0) | 0.04 |
|  | 40-49 | 7,953 (17.3) | 31 (5.7) | 0.39 |
|  | 50-59 | 6,624 (14.4) | 51 (9.4) | 0.77 |
|  | 60-69 | 3,748 (8.1) | 92 (16.9) | 2.45 |
|  | 70-79 | 1,785 (3.9) | 152 (27.9) | 8.52 |
|  | 80 and above | 1,108 (2.4) | 207 (38.1) | 18.68 |

Figure 1A presents the gender proportion of reported cases by age groups and cumulative morbidity ratio by gender and age group. Across age groups, the proportion of male cases is higher than 50%, except for those aged 80 years and above (χ^2^ test, p-value <0.0001). Cumulative morbidity ratio by gender and age group is presented in Figure 1B, indicating that ratio among males is higher than among females across age groups (proportion test, p-value < 0.000) except for those aged 80 years and above (proportion test, p-value = 0.13).

**Figure 1.**
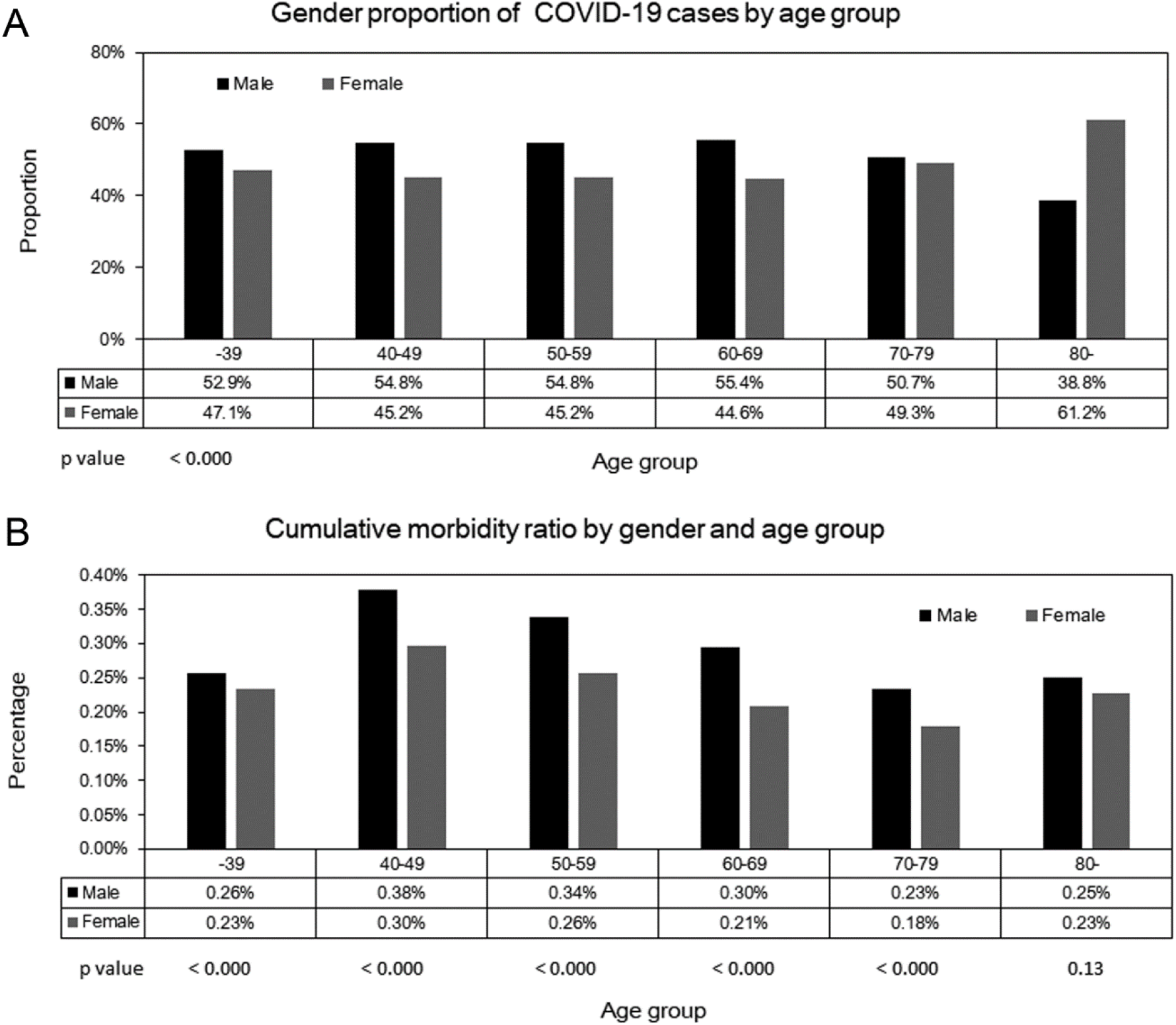
Gender proportion of reported COVID-19 cases by age groups (A), and cumulative morbidity ratio by gender and age group (B), March through May 18^th^, 2020, Chile

Figure 2 displays the cumulative cases of CoVID-19 by age group (A through G), and the cumulative deaths by age group (H through N) over time. The figure suggests cumulative cases of CoVID-19 are growing faster than cumulative deaths. The growth curve for cumulative cases across all age groups and for persons aged 0-39 appears to increase exponentially after around day 30 (April 30^th^, 2020), while exponential growth in cumulative deaths for all age groups appears to occur after day 45 (May 15^th^, 2020).

**Figure 2:**
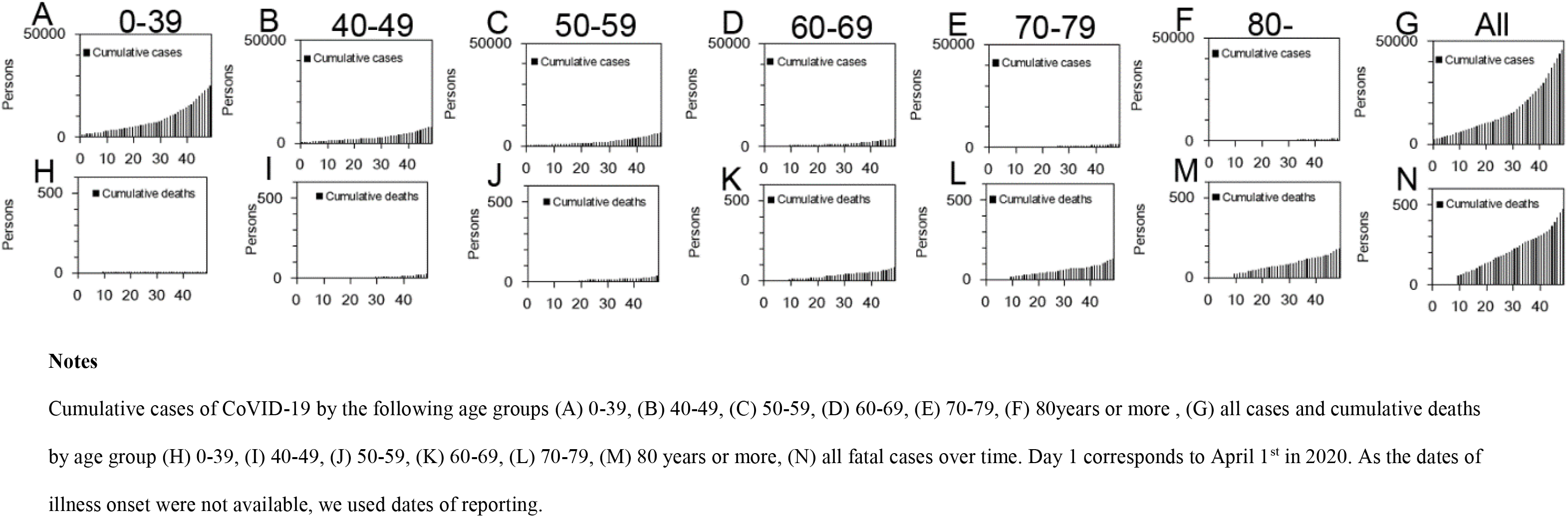
Temporal distribution of cases and deaths by age group due to COVID-19, March through May 18th, 2020, Chile.

Figure 3 shows observed and model-based posterior estimates of the crude CFR of CoVID-19 by age group (A-G) and time-delay adjusted CFR by age group (H-N). Black dots show crude case fatality ratios, and light and dark indicate 95% and 50% credible intervals for posterior estimates, respectively.

**Figure 3:**
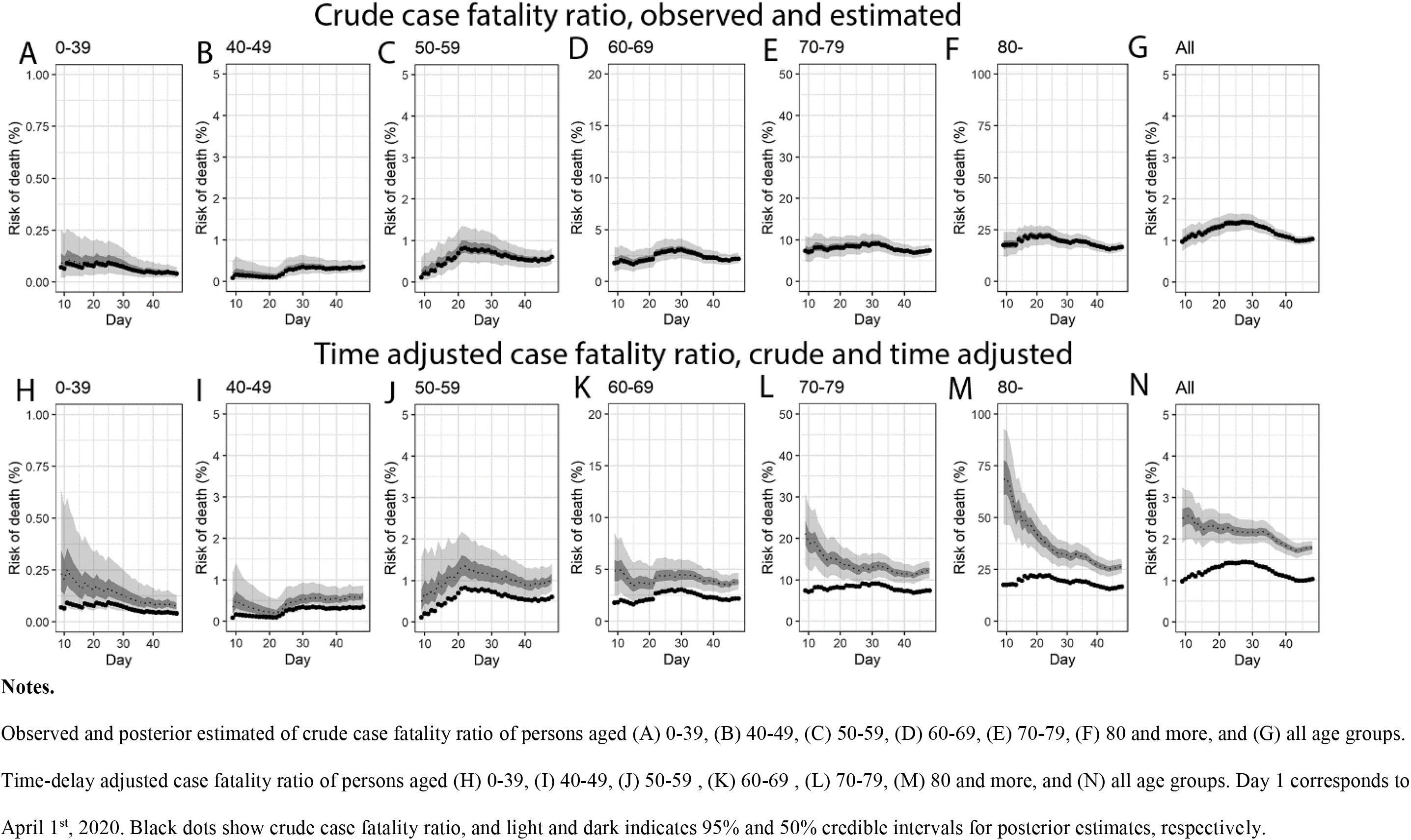
Temporal variation of risk of death caused by COVID-19, March through May 18^th^, 2020, Chile.

Overall, our model-based crude CFR fitted the observed data well. Crude CFR for all age groups (G) increased at an early stage of the epidemic, peaking around April 25^th^ and was followed by a decrease in CFR for about two weeks. There is a suggestive increasing trend across most age groups starting around May 10^th^, 2020. Our model-based posterior estimates for the time-delay adjusted CFR are substantially higher than the crude observed CFR. The overall adjusted CFR follows a decreasing trend, except for infected patients aged 50-59 years, where the adjusted CFR increases until around April 21^st^.

The most recent estimates (May 18, 2020) of the time-delay adjusted CFR are 0.08% (95% CrI: 0.04-0.13%) for persons aged 0-39, 0.61% (95%CrI: 0.41-0.87%) for those aged 40-49, 1.06% (95%CrI: 0.76-1.40%) for those aged 50-59, 3.79% (95%CrI: 3.04-4.66%) for those aged 60-69, 12.22% (95%CrI: 10.40-14.38%) for those aged 70-79, and 26.27% (95%CrI: 22.95-2980%) for persons aged 80 and over. The overall time-delay adjusted CFR is 1.78% (95%CrI: 1.63-1.95%) across all age groups (Table 2, Figure 4).

**Table 2.**
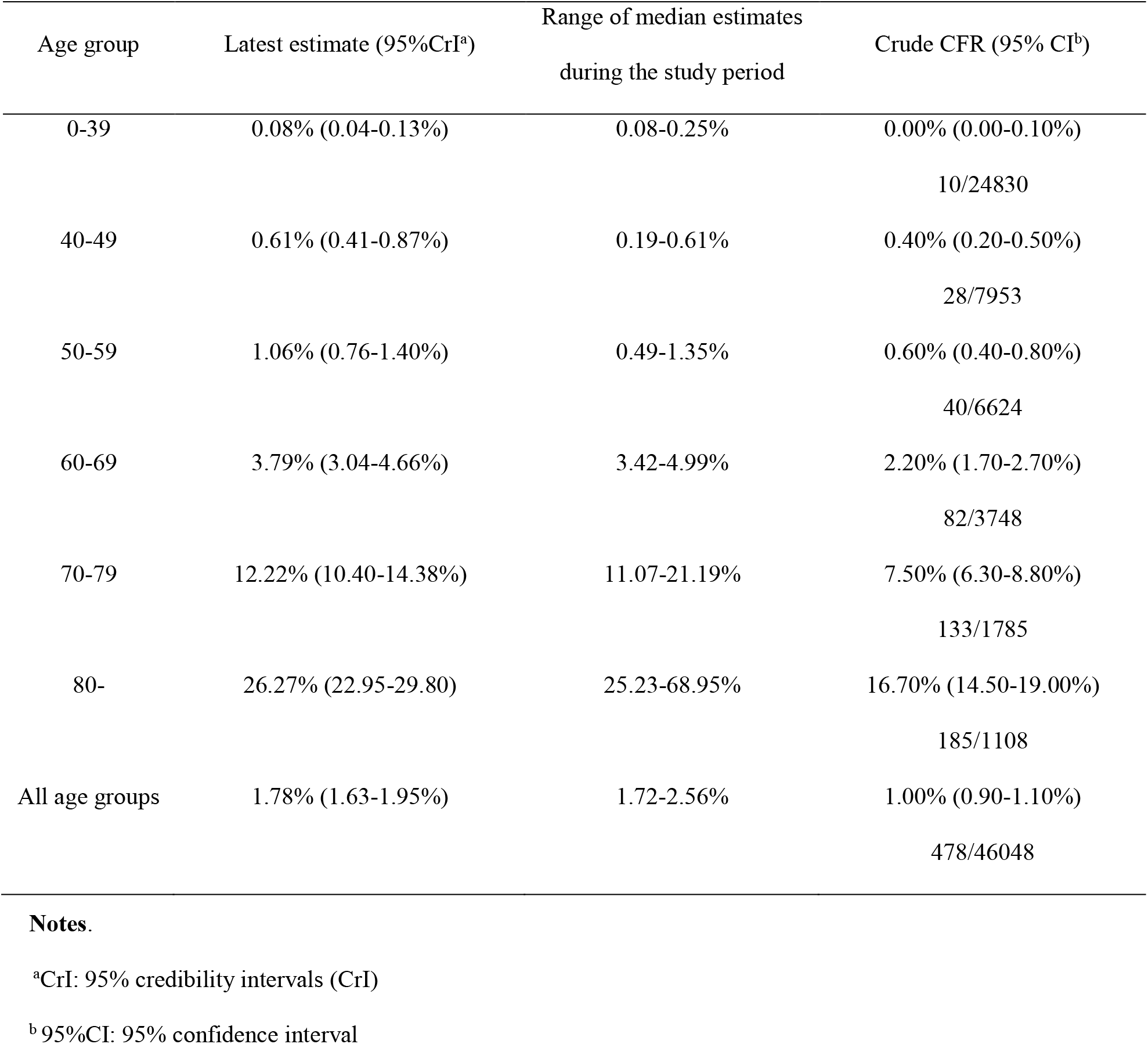
Summary results of time-delay adjusted case fatality ratio of COVID-19 by age group in Chile, 2020 (May 18, 2020)

**Figure 4.**
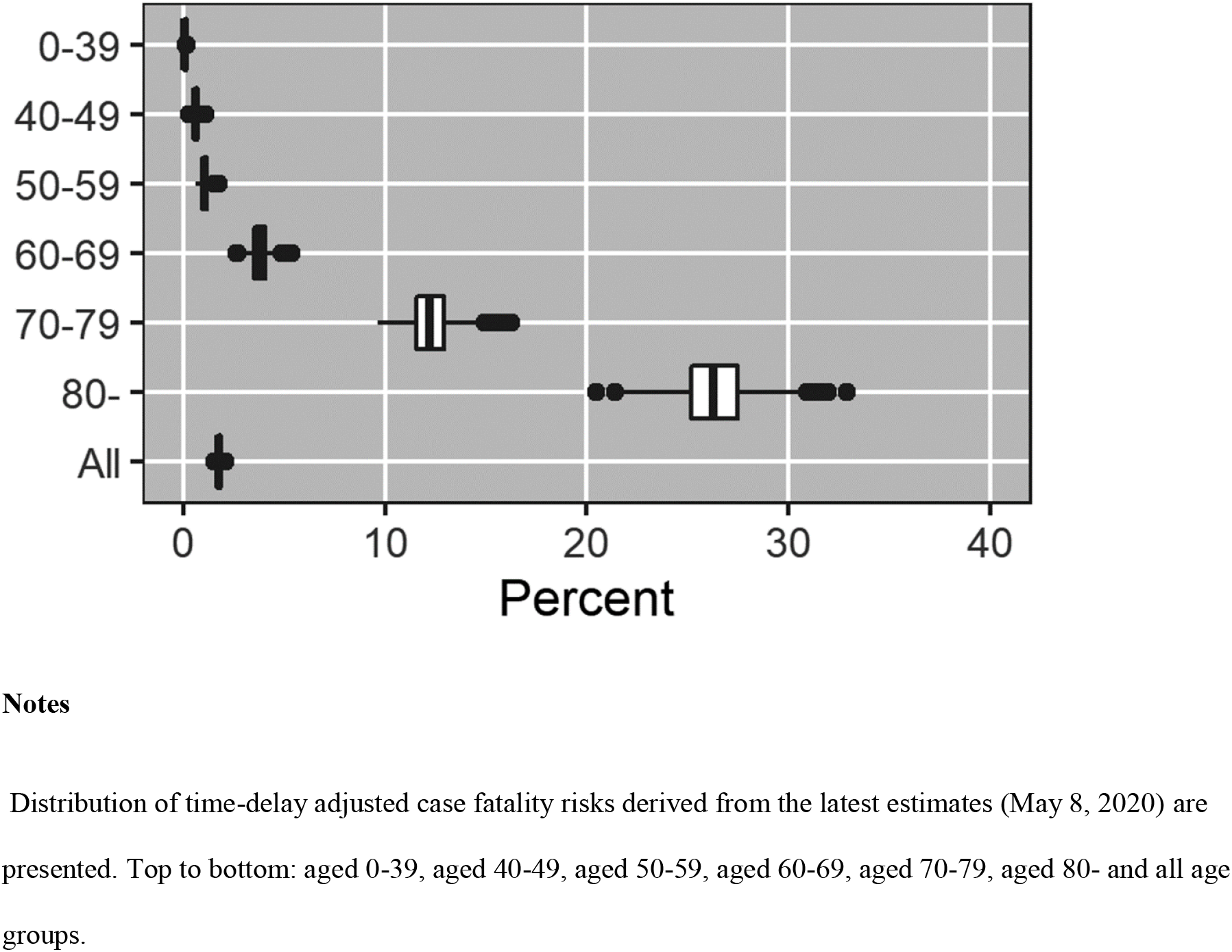
Latest estimates of time-delay adjusted risk of death caused by COVID-19 by age 421 group, March-May 2020, Chile.

## Discussion

To the best of our knowledge, this is the first study to estimate the time delay adjusted CFR by age group for COVID-19 in Latin America, a region that has yet received little attention during the ongoing coronavirus pandemic. Consistent with other recent COVID-19 research [23, 25, 40], our results show the COVID-19 epidemic in Chile disproportionately influenced seniors, especially those aged ≥ 70 years. These results suggest that an aging population could exacerbate the fatality impact of COVID-19 [41], similar to influenza and respiratory syncytial virus [42], and consistent with data available from Italy[41]. The comparatively low CFR observed in Chile during the early stages of the epidemic [17], probably reflected the age structure and socioeconomic status of initial cases. During the first weeks of the pandemic, COVID-19 cases occurred among relatively young age groups, with most transmission occurring among individuals between 20 and 60 years of age, and in high-income communities with better access to healthcare and lower prevalence of risk factors for severe COVID-19 [12, 43, 44].

Our latest estimates (as of May 18^th^, 2020) of the adjusted CFR among those aged 80 and over reach values as high as 26.3% (95%CrI: 23.0-29.8), an estimate that is 328-fold higher than our estimates for those aged 0-39 (0.08%), and 2.2-fold higher than our estimates for those aged 70-79 (12.2%).

An upward trend in the crude CFR, as seen for the overall population through most of April and particularly for the 50-59 age group, suggests the transmission is spreading to more vulnerable populations. Interestingly, throughout March, most of transmission occurred among relatively young, better-off populations. In Santiago, which has more than 75% of COVID-19 cases in Chile, targeted enforced quarantines were put in place starting March 28^th^ through mid-April in seven municipalities, six of which are among the richest in Chile [45]. Epidemic transmission thus moved during April towards lower income municipalities, where social distancing measures are more challenging to comply with, due to a higher proportion of the population participating in the informal economy, higher population density, poorer infrastructure, and lower quality healthcare [12, 46]. An upward trend in the crude CFR could also result from an increasing number of unreported cases due to saturated testing capacity. However, this is an unlikely explanation as RT-PCR testing capacity has increased with cases, maintaining an average positivity rate (positive tests / total tests) of 9.7% throughout April (SD: 2.4%). However, results show an increase in crude CFR around day 45 (May 15^th^, 2020), probably reflecting the exponential increase of cumulative cases around day 30 (April 30, 2020), and a substantial increase in positive test rate (average in May 1-18: 15.2% SD:4.5), which suggests there is probably an increasing proportion of under diagnosed COVID-19 cases in Chile [47].

The downward trend in the adjusted CFR at the early stage of the study period may have been influenced by reporting delays. In particular, the observed differences in our estimates of the crude and adjusted CFR are directly due to the time-delay which we assume fixed during the course of the epidemic.

The small proportion of men (38.8%) among CoVID-19 cases in people aged 80 and over is probably attributable to the relatively small male population size for that age group; men represent only 37% of the population >80 in Chile [48]. Higher mortality among men has been reported in China and the U.S. [49, 50], our data provide the opportunity to examine the CFR by gender and age.

Our study is not exempted from limitations. Our CFR estimates are probably affected by under ascertainment, as has been estimated elsewhere [24-28], which may have pushed our estimates upwards. Infectious disease with a substantial share of asymptomatic or mild infections, such as COVID-19, may be more accurately characterized by infection-fatality risk (deaths / infected people), but those data are not yet available in Chile.

In conclusion, using real-time epidemiological data from a high COVID-19 testing setting in Latin America, we found that COVID-19 epidemic in Chile disproportionately affected seniors, especially those aged ≥ 70 years, suggesting an older population could exacerbate the death toll brought by COVID-19. COVID-19 is already imposing a high death toll in Latin America. Case fatality rates in Chile suggest the health system is not yet overwhelmed, but the epidemic is expanding fast, and healthcare demand may soon exceed capacity. These real-time estimates may help inform decisions by public health officials in the region, and underscore the need for continuous efforts to face this pandemic.

## Data Availability

All data used in this study are publicly available in the webpages of the Ministry of Health and the Ministry of Science in Chile.

http://www.minciencia.gob.cl/covid19

https://gisanddata.maps.arcgis.com/apps/opsdashboard/index.html#/bda7594740fd40299423467b48e9ecf6

## Acknowledgments

This work was supported by the Japan Society for the Promotion of Science (JSPS) KAKENHI “[grant Number 20H03940]”; the Leading Initiative for Excellent Young Researchers from the Ministry of Education, Culture, Sport, Science & Technology of Japan to [KM]; National Science Foundation NSF “[grant 1414374]” as part of the joint NSF-National Institutes of Health NIH-United States Department of Agriculture USDA Ecology and Evolution of Infectious Diseases program; UK Biotechnology and Biological Sciences Research Council “[grant BB/M008894/1]” to [GC]; and the Millennium Science Initiative of the Ministry of Economy, Development, and Tourism, Government of Chile, “[grant Millennium Initiative for Collaborative Research in Bacterial Resistance]”.

